# “It’s possibly made us feel a little more alienated”: How people from ethnic minority communities conceptualise COVID-19 and its influence on engagement with testing

**DOI:** 10.1101/2021.04.06.21254961

**Authors:** Tushna Vandrevala, Lailah Alidu, Jane Hendy, Shuja Shafi, Aftab Ala

## Abstract

**Objectives:** The cultural beliefs, practices and experiences of ethnic minority groups, alongside structural inequalities and the political economy play a critical, but overlooked role in health promotion. The current study aims to address this deficit; understanding how these groups conceptualise COVID-19, and how this influences engagement in testing, with the future aim of developing targeted communications to address the challenges of testing uptake.

**Method:** Black (African and Caribbean) and South Asian (Indian, Pakistani and Bangladeshi) community members were purposefully recruited across the UK. Fifty-seven semi-structured interviews were conducted and analysed using principles of Grounded Theory.

**Results:** The findings illustrate that Black and South Asian’s conceptualise COVID-19 as a disease that makes them visible to others outside their community; in having more severe risk and suffering worst consequences; resulting in fear, stigmatisation and alienation. Views about COVID-19 were embedded in cultural beliefs, relating to culturally specific ideas around disease, such as ill-health being God’s will. Challenges brought about by the pandemic were conceptualised as one of many struggles, with the saliency of the virus contextualised against life experiences. These themes and others influenced engagement with COVID-19 testing. Testing was *less* about accessing timely and effective treatment for themselves, and more about acting to protect the family and community. Testing symbolised a loss of income, anxiety and isolation, accentuated by issues of mistrust of the system, and not being valued, or being treated unfairly.

**Conclusion:** In tackling these challenges, we conclude that health communications should focus on counterbalancing the mistrust, alienation and stigmatisation that act as barriers to testing, with trust built using local credible sources.

## Introduction

The global COVID-19 pandemic has highlighted the scale of health and social disparities, the complex interaction of socioeconomic disadvantage, influencing exposure to SARS-CoV-2 (from now referred to COVID-19), which has profound consequences for ethnic minority groups. In order to develop a more targeted and inclusive public health response we need a better understanding how ethnic minority groups (EMGs) conceptualise COVID-19, and how this conceptualisation influences engagement in testing,

The COVID-19 pandemic is considered to discriminate, inflicting a disproportionate burden of illness and death across BAME communities in the UK^1,2^ and a disproportionate mortality among BAME healthcare workers.^3^ Individuals from different ethnic backgrounds vary in behaviours, co-morbidities, immune profiles and risk of infection.^4^ EMG may have increased incidence of co-morbidities (insulin resistance, diabetes mellitus, cardiovascular disease, central obesity and hypertension, vitamin D deficiency) with these conditions linked to poor outcomes for COVID-19.^5^ Material deprivation, the location of residence, household composition, overcrowding in housing, high-risk occupation, comorbidities, and limited knowledge of the healthcare system leads to unequal access to treatment.^6^ EMGs are more likely to live in shared, sub-standard or overcrowded accommodation, with extended family in multi-generational households, with implications for transmission from younger to older and more vulnerable household members,^3,7^ associated with higher positivity rates for COVID-19.^8^ Lower English language proficiency, poorer transportation, and lack of recourse to public funds are additional contributory factors.^4^

Ethnic minorities make up a significant proportion of front-line public-facing jobs, (healthcare, social work, hospitality, retail, delivery, household services), and in menial jobs that can place them at greater risk of exposure.^9^ They are more economically vulnerable to the current crisis; working in shut-down sectors, experiencing higher partner unemployment and lower potential for buffering income or savings and fall outside of government safety nets.^10^ Low□income workers face financial disincentives for absence when they are sick or vulnerable, increasing workplace transmission and highlighting the difficulties in engaging in testing and self-isolating.^11^

Migrants may be late presenters to health services^12^ and face multiple barriers^13^, presenting only where necessary due to concerns around immigration status, mistrust, lack of knowledge of the health system, and poor access. EMGs and migrants can lack knowledge and awareness of COVID-19, putting them at risk of the spread of misinformation^7^. Perceptions of risk regarding testing for communicable diseases may be fundamentally different from western beliefs, resulting in people inadvertently putting themselves at greater risk.^14^ Fatalism, belief in God and illness being attributed to external factors, focus on familial risk rather than individual risk can as barriers to testing for communicable diseases.^14^ There is low awareness of the danger of the pandemic, especially among migrants from sub-Saharan Africa,^15^ combined with misconceptions regarding the prevention of COVID-19 infection, and the use traditional remedies.^16^

COVID-19 incidence and testing uptake by migrant status account for a disproportionately high number of COVID-19 infections combined with low rates of testing.^8,17^ While testing and treatment for COVID-19 are free of charge, concerns remain that these exemptions do not mitigate barriers. Migrants are concerned that COVID-19 treatment is chargeable, or that seeking care or putting themselves forward for testing could mean losing their job and/or be deported.^11^

Psychological health behaviour models and public health models usually focus on individual level factors that predict intention to engage in health protecting behaviours, such as testing and do not specifically account for the differing experiences of minority groups.^18^ The conceptualisation of health behaviour as being the result of an individual decision-making process does not account for sociocultural and socio-structural factors, and the importance of social connectedness and integration.^18^ Public health messages can fail to have the necessary impact, with a need for culturally specific, targeted interventions. This research attempts to address this gap by specifically asking how EMGs conceptualise and understand COVID-19, taking into account social, cultural and lived experiences, and how these understanding impact on willingness to test (Phase 1 of the project, reported here). In phase 2, we will develop communications in the form of evidence base films and guidance documents to address these challenges and offer solutions, embedded within cultural norms and delivered by local community champions and trusted advocates, with the EMG feeling the messages delivered are for them and about them.^19^

## METHOD

### Study Design

The study is qualitative, underpinned by a constructivist perspective, in which participants are viewed as creators of their social world, which the researchers subjectively interpret.^20^ The research is located within a ‘big Q’ approach to qualitative work; a paradigm that does not seek to quantify not aspire ‘objectivity’.^21^ Instead, meanings and interpretations participants attach to their own experiences are acknowledged and grounded in specific social, historical and cultural contexts.

### Recruitment and participants

After approval from the author’s institutional ethics committee, initial contact was made with key individuals within community hubs across England; such as charities and community organisations, faith organisations, and women’s centres with high levels of attendance by Black and South Asians. We have researched previously in a similar area, so recruitment was facilitated by these individuals, our existing contacts and our wider project partners. The researchers who ran the interviews and analysed the data were integrally involved in each stage. They are members of UK Black (African and Caribbean) or South Asian (Pakistani, Bangladeshi, Indian) communities and therefore “insider researchers,” sharing common knowledge, a common identity and some common life experiences. This commonality facilitates meanings within our participants’ word.^22^

Participants and stakeholders were recruited nationally from a range of socio-economic backgrounds, with differing cultural, philosophical and religious background and different occupations, ages and health status. Stakeholders were defined as people embedded within the ethnic community groups above (and sharing the same ethnicity) and working with communities either formally (public health and allied health professionals) or informally (community leaders, faith leaders, advocates).

Eligibility criteria were being 18 years of age or older, from a Black (African or Caribbean), and South Asian (Pakistani, Bangladeshi, Indian) background, ability to comprehend and speak English to a sufficient level in order to understand instructions; ability to sign a consent form; and availability to be interviewed via MS Team, Zoom, Skype or phone call. First and Second-generation migrants were included. The research team met regularly during fieldwork to review progress and ensure that a diverse range of people were recruited. Twenty-eight participants identified themselves as Black (African and Caribbean) and twenty-nine from a South Asian background (Indian, Pakistani and Bangladeshi) (see Table 1).

### Data Collection

Semi-structures interviews were chosen to ensure that core questions were asked of all participants while providing scope to explore relevant but unanticipated domains of experience and reflection. Interviews were conducted between August and December 2020 (lasting 45-80 min), by a researcher from the African community who was culturally and linguistically equipped to understand the target population via Zoom/MS Teams (n=47) and telephone (n=10). Participants were given a £25 amazon voucher for their time.

The research team developed the interview guide; a list of topic areas using open-ended questions and prompts, which was frequently annotated/moderated during the progression of the study.^20^ The interview guide explored perceptions in which the coronavirus pandemic has affected/influenced respondents and members of their communities. The questions iteratively developed as ideas emerged and addressed issues relating to how participants conceptualised the pandemic in their lives. We explored the impact of the disease, the effectiveness of guidelines and messaging, and facilitators and barriers to engaging with various health protecting behaviours (such as hand washing, social distancing, social isolating and engaging with testing). In this paper, we specifically report data relating to their willingness to engage with testing for COVID-19.

After 57 interviews, the research team agreed that data saturation had been achieved, with no new significant insights emerging. The recordings were transcribed verbatim. Further methodological details are available from the authors.

### Data Analysis

Our analysis was based on the principles of grounded theory, following distinct first-order and second-order phases.^23^ The first phase involved gaining a familiarity and understanding of the literature and context on health inequalities and COVID-19. Following a review of the literature, two authors conducted an in-depth reading of the interview transcripts and recorded observations about the participant’s experiences in the form of ‘memos’ and reflections of the potential relationships, structures and attitudinal responses. In the first stage of data structure, we (a) used an axial open coding approach to inductively code the lived experience of participants, (b) our axial coding grouped together similar codes to form categories^23^ and (c) theorised and contextualised the framing (see table 2 for the results of this data structuring process). This first-order phase of data analysis began after the first interviews and continued at regular intervals. At many times our phases overlapped, and we moved back to first-order phase activity on a number of occasions, to verify and check emerging findings.

For the second stage, presented below, we drew on a complex, intertwined relational and dynamic contextual, psychological and societal interactions to gain theoretical insight into why some participants seemed unable or unwilling to seek testing, and what issues contributed to this. We organised our findings into three different contextual levels: the individual, the familial and a system/societal lens (specifically drawing on the analytic work for this latter lens). ^23^

The process of analysis was made transparent by conducting credibility checks and optimising coherence. The consolidated criteria for reporting qualitative studies (COREQ) were followed.

## Results

### COVID exacerbates visibility and stigma

Participants revealed that the narrative adopted by the media and the UK Government in highlighting the disproportionate impact of COVID-19 on EMGs made others outside their community aware of their contributions to society as frontline workers, working in the NHS and other important and essential caring services.

> *It changed the way that other people saw us. I don’t think that it changed the way we saw us. We knew that we were working hard at the butt end of racism. We knew that it is members of our community who were supporting the NHS, keeping the carer infrastructure alive in this country. We knew all of these things about ourselves. What happened with COVID, is it forced other people who didn’t see us, to see us, all of a sudden*. (Jamaican, Male, 58)

The majority of our participants expressed that the new visibility highlighted socio-economic and health inequalities that already existed, and in the current pandemic manifested into disproportionate health outcomes for EMGs. Participants were acutely aware that their previous health conditions, co-morbidities, their occupations (increased exposure), their living arrangements (i.e., multi-generational households) and cultural values and norms put them at “additional risk” of adverse outcomes, this led to fear, anxiety and loss.

> *I work in the hospital - it is really frightening knowing that you are going into work and you don’t know what to expect* (Zimbabwean, Female, 26)
>
> *I think minorities are the ones that are working in the frontlines or doing the jobs that people don’t want to do. Then I guess I’m more at risk in a sense*. (Djibouti, Female, 19)
>
> *We are mostly, I would say in service sector*.. *more or less front-line staff, working on buses, trains or catering industry or taxis or in hospitals. You are more likely to get infected*. (Indian, Male, 31)

Overall, our participants expressed that they did not feel supported by the healthcare system, in terms of helping to mitigate this “risk”. Majority of participants said that the COVID-19 guidance and messaging was not about them, or for them, and instead favoured a ‘white’ privileged position that did not reflect their reality.

> *Yeah, a number of these measures are kind of very much aligned to a particular category of people to work from home and not many people have that benefit or privilege to work from home. I think it’s a white collared mentality to feel that everybody would be able to work from home. It becomes a challenge for those who work outside their home to be able to do it and have an income. Most immigrant families aren’t that privileged. Most of us are in dirty, demeaning or deadly jobs*. (Jamaican, Female, 61)

Black and South Asian participants believed that the discourse around being “disproportionate represented” also implied being “disproportionately responsible” for the spread of the virus, and they were in some ways being “blamed” for spreading COVID which increased alienation and stigma.

> *I think somehow there’s always this narrative that we’re the other. COVID, obviously there’s research and statistics showing that our communities have been affected far more greater than white counterparts. And so, that’s another problem to us, if that makes sense… We have COVID, and nobody else seems to have it. And we’re the ones that are spreading it around* (Pakistan, Female, 22)

Therefore, participants expressed that COVID made them visible to others outside their community, but this was often attached to blame, stigma and othering, which contributed to a sense of growing alienation.

### Self-efficacy an antidote to fear and stigma

COVID is embedded in cultural beliefs about illness amongst EMGs, where illness was seen as a sign of weakness or vulnerability and participants were aware of the stigma that a diagnosis of COVID-19 could bring:

> *I think there’s also a sense of stigma in the Somali community, when it comes to illness and diseases, people don’t want to be seen as weak or vulnerable. So, I think there’s also that mentality of just firming it and hoping for the best, and not wanting to be seen as suffering from COVID-19*. (Somali, Male, 27)

In some cultures, illness was considered as a sign from, or the will of God, with such attributes to illness meaning it could not be avoided or eradicated unless God decided, with people having little or no contribution to the impacts or outcomes of the pandemic.

*We are deeply religious people, and they say, whatever you behave, will be. If it’s your time, it’s your time*. (Somali, Female, 57)

> *People say Allah knows why this is happening*. (Bangladeshi, Male, 65)
>
> This fatalism can lead to a lack of preventative behaviours and self-efficacy. The sense of lacking any agency was further intensified by the EMGs misconceptions about COVID-19; namely that “*people back home not getting it! We are immune*”. This strong narrative was accompanied by theories that God was intervening. Fate, destiny, faith and prayer were offered as strategies for protection from the effects of the pandemic.
>
> *The pastor would encourage us to pray that nobody is going to die of Coronavirus and we return back to church safe and alive*. (Ghanaian, Male, 35)
>
> *Coronavirus is punishment from God for disobedience*. (Pakistani, Female, 45)

A lack of agency and self-efficacy reflected a lack of willingness to engage in testing for themselves. Perhaps the most surprising and unexpected finding was that our participants (both Black and South Asian) did not conceptualise testing as a means of seeking timely treatment to protect themselves. The main motivation to test for the virus was to avoid the spread to other family members or their community. Both Black and the Asian communities have strong family ties and the need to protect the family was a strong social norm. This salient point was often expressed as strong motivator for the community to participate in testing and other protective behaviours.

> *You know Africans have always been family, family is really important…*.*you don’t want to infect anyone* (Cameroonian, Female, 58)
>
> *I do not want to put them at risk because me being young might be able to cope better and they might not be able to cope as good as I would*. (Indian, Female, 23)
>
> It was evident in the data that high levels of fear and anxiety around COVID-19 its impact, led to a lack of self-efficacy to engage with the process of testing, with fear of catching COVID-19 at a testing centre salient:
>
> *Because I do feel as though there’s probably a level of stigma (to go to the testing centre). I even heard one person telling me, even by going to the test centre, you’re increasing your likelihood of getting coronavirus, because everyone there probably has coronavirus. So, even though they had symptoms, they were very reluctant to go, because they were worried that they might not have it, but by exposing themselves to people at the test centre that have it, that they themselves will catch it*. (Ivorian, Female, 29)

Majority of participants also expressed they lacked the necessary knowledge and skills required to access the testing-booking system, with limited IT skills and access to smart devices.

### Struggles and the burden of being a migrant

For majority of our participants the COVID-19 pandemic was viewed with a lens of “one of many battles and struggles” they face and therefore, had a less important saliency than expected. In some cases, the COVID virus was constructed as a “small and inconsequential” virus, in terms of surviving different forms of hardship.

> *I hear even some people were saying, we came through civil war, this is nothing for us*. (Somali, Male, 27)

People from the African continent suggested that COVID was:

> *It’s just a disease, people are dying more from other illnesses, why are we complaining*. (Cameroonian, Female, 58)

Participants were often more concerned about the economic consequences of the pandemic than falling ill from virus. Providing for their families was the most important “duty” and testing and having to isolate as a result was not considered important or relevant to those who needed to worry about providing.

> *People are worried not just about Covid-19 they are worried about their last meal. Some people will not want to say they have Covid-19 because they have to isolate. That means less income* (Nigerian, Female, 45)

In EMGs, the man is often perceived as the main bread winner/provider for the family. Where incomes are uncertain, and the threat of job losses very tangible, the loss of income from having to isolate and the financial burden of this acted as a disincentive towards testing. The struggle to provide basic needs for subsistence outweighed the need to strive for safety (for something which might or might not happen). Perceptions of testing were closely tied to views on isolating.

> *If the man is the breadwinner within the household, financial circumstances as well would be a problem. The financial burden is a problem. If we have to isolate ourselves for 14 days or whatever days, that’s two weeks of wages. And who’s going to support that? And I think that probably is one of the key factors as well*. (Bangladeshi, Male, 36)
>
> *It’s because the implication of having a positive test and isolating, as well as the people around you. If you are the breadwinner, then it’s going to be difficult, isn’t it, to isolate and not work and get paid. And there is also a proportion of our people not in high skilled jobs, illegal immigrants, not having the right to stay, and all that, are likely to play down their symptoms, and not go for tests and isolate, because then, how is food going to be put on the table for the family?* (Ghanaian, Female, 58)

### Genesis of mistrust and its impact

Participants expressed that the health system did not understand them or welcome them.

There was fear and anxiety about falling sick and being treated poorly by the NHS.

> *If I get sick what will my situation be? What is the priority. That is what I think, why am I less privileged or will I be equally treated the way other ethnic groups are treated. I am very anxious. I don’t want to get sick*. (St Lucia, Male, 40)
>
> *They think that if you end up in hospital, you will be dead. There are some people who think they would actually deliberately kill you if you go to hospital*…*You aren’t prioritised. They would just let you die*. (Indian, Male, 83)

Majority of participants drew on their previous experiences to conclude that the Government and the healthcare system had failed to prioritise their health, and the health needs of their community, with historical issues of mistrust brought to the fore during the pandemic. Participants were likely to avoid booking a test because of privacy concerns. They said that the lengthy questions asked before one could book a test and the information required would be used to monitor them, track their immigration status and would be shared with the home office, resulting in deportation.

> *I think to motivate people to go for tests, it has also come with some assurance that look, this is not going to expose anyone to the Home Affairs or anything like that. And the essence of the healthcare team is not to expose anyone to immigration or something like that* (Ghanaian, Female, 58)
>
> Their suspicions and mistrust about testing permeated to the test and trace app – which was perceived as a system to monitor their lives.
>
> *I don’t know one person that tested as track and trace. This tracing is it just people in general rather than with Covid. It’s all control, really*. (Barbados, Female, 59)
>
> Majority of participants expressed that the Government had intentionally brought about rules and guidelines without considering their specific needs.
>
> *But I feel the government has let us down right from the beginning. It’s not just for the COVID. Even before COVID, the health centres, doctors were not given much guidance about people’s health in general*. (Bangladeshi, Female, 48)
>
> *It was literally introduced in the evening before Eid and everybody was feeling really like, they’ve just done this because it’s Eid tomorrow and they don’t want us to mix. And then the policing, I think they’ve always felt overly policed, as it is. And now it’s like, ethnic minorities have bigger families and they’re more likely to be breaking the rules. And people are getting reported and I think they feel overpoliced in that regard*. (Pakistan, Female, 30)

Surprisingly, none of our 57 participants suggested that they would consider COVID-19 testing as a means to access timely and effective treatment. What is evident from our data is that Government messaging around testing (and its focus on symptomatic testing) discouraged our participants from putting themselves forward for testing.

> *I guess because the government, especially the NHS, sorry, because they’ve made it so clear to only get tested if you have symptoms, and because of their symptom list some people don’t fit into that. So they don’t want to waste the NHS money and time and go and get tested. I am one of those people that will not go to get tested probably as well unless I actually felt I was on my last legs or something like that, to me it feels like a waste of NHS time and money as well*. (Pakistani, Female, 24)

## Discussion

This study aimed to understand how EMGs conceptualise and understand COVID-19, taking into account social, cultural and lived experiences and how these understanding influence willingness to test, with the future aim of developing communications that address these challenges and offer solutions. It is crucial that public health professionals understanding EMG perceptions, needs and concerns related to prevention and testing of COVID-19 so we can design and implement specific, culturally relevant, useful and trusted interventions. The landscape of COVID-19 testing and uptake are changed at incredible pace, with the focus now also shifting to asymptomatic (mass) testing. However, the results from this study are likely to remain relevant across the changing landscape as it demonstrates that understandings of COVID-19 are embedded in cultural beliefs, economic concerns and long standing issues, which are not transient.

Our findings highlight that people from EMGs expressed that the media emphasis on the disproportionate representation on COVID-19 mortality and illness made them more visible. This visibility stigmatised them, as they felt blamed for COVID-19-related exposure, mortality and spread. Furthermore, the primary concerns for Black and South Asian people were to provide for their family, rather than the threat of COVID-19, and this impacted on their engagement with testing. Participants talked about being “forced to self-isolate” if they had severe symptoms. Isolation was perceived as something they were not able to do rather than something they were unwilling to do. They would be unable to isolate due to their immediate need to provide for their families, hence, it was easier not to engage in testing.

Cultural and religious beliefs about illness and COVID-19 and misconceptions about COVID-19 self-efficacy led to high levels of anxiety, fear and loss. Perhaps, the most salient finding was the lack of individual self-efficacy among participants, with not a single participant expressing that they would access COVID-19 testing to access timely and effective treatment for themselves. For our participants, COVID-19 was embedded in a political system of mistrust. COVID-19 messages reinforced historical levels of racism, discrimination and mistrust.

Our findings draw attention to perceived ‘narratives of blame’ used by politicians and media, with people from EMG reporting being stigmatised and alienated during the COVID-19 pandemic.^24^ Where concerted efforts to control and prevent the risk were required of everyone, EMG perceived they were positioned to pose an extra risk to society and were therefore more responsible for the spread of the virus. The emphasis on race as a risk factor, portrayed in media and by Government and policy makers appeared to exacerbate the blame, stigma and alienation experienced by our participants, suggesting communications need to place less emphasis on race or ethnicity and explore wider social determinants of health.

The current testing system does not take enough account of the complexity many minorities face (such as loss of income and the struggle of living on the poverty line) when asked to test. Unless government policy provides material support (in the form of income and sufficient space) for people to self-isolate, many EMGs will not adhere to self-isolating protocols due to practical issues, such as having to feed one’s family, rather than any lack of motivation. ^24^ As we move into the next stages of the pandemic, even more emphasis is likely on both symptomatic and mass (asymptomatic) testing, so there is an urgent need to avoid viewing testing and self-isolating as separate protocols and work to fill the gaps our participants have identified.

When faced with the threat of stigma and fear as a result of a global pandemic, individuals are likely to consider psychological resources at their disposal, such as self-efficacy; the belief that one has the ability to act to manage a potential threat.^25^ Behavioural change theories have consistently emphasised the importance of self-efficacy, without putting needed emphasis on community-efficacy or familial-efficacy beliefs. Our study reveals participants were motivated to test to protect their families and their communities, in contrast to protecting themselves. This strongly suggests a need to change emphasis from individual self-efficacy to collective efficacy (pushing the belief that groups, can work together to achieve an intended outcome).^26^

One of our unique findings was that for EMGs COVID-19 was embedded in a political system of mistrust, and the strength of this mistrust across the healthcare. Disease outbreaks and pandemics can expose and exacerbate long-standing health inequities, systemic racism and marginalisation. Recent studies from the US have highlighted similar issues of mistrust in healthcare and government among African Americans,^27^ with decades of racism, marginalisation and discrimination isolated communities and making them particularly vulnerable and susceptible to COVID-19.^28,29^ Our work is the first to emphasise the impacts of this historical mistrust on the UK test, trace and isolate systems. It is vital that consistent efforts are made to work with community champions and leaders during and beyond the pandemic, to start to shift and reframe beliefs, towards a more positive narrative that positions institutions such as the NHS as being equally welcoming and there for everybody.

Previous psychological theories of behavioural change and public health models usually focus on individual level factors that predict intention to engage in health protecting behaviours, such as testing. These models fail to account for the granularity of sociocultural and socio-structural determinants, which our study has highlighted as salient for COVID-19 related health-promoting behaviours. Our study shows that concern for economic consequences is important in predicting protective health behaviours and that efficacy, at an individualistic level is not particular useful. Much traditional theoretical framing of behavioural change fails to appropriately focus on the power of the family and collective community and fails to fully appreciate the influence of these factors (both positive or negative).^30^ Our work clearly shows the role of social connectedness and integration^18^ and the systemic factors that are of particular relevance to EMG and migrant groups. Therefore, there is a need to develop pragmatic, culturally embedded models of health behaviour that provide complex, multi-layered contextual, cultural and socioeconomic granularity, specifically relevant to living with this modern-day pandemic.

Test, trace and isolate systems are seen by public health and government officials as fast and effective ways to control the spread of the virus. Our study suggests this not the case in EMGs. Firstly, in tackling this misalignment we need to focus on communications that counterbalance mistrust, alienation and stigmatisation, and work to actively built faith in these methods using local trusted sources. There is a need for access to clear, accurate, targeted, and visible educational materials from authentic and trusted local sources. We need to help counterbalance mistrust in NHS by facilitating and encouraging the community to talk more openly about COVID-19. Secondly, the messages on COVID-19 should focus on building familial and community efficacy beliefs, harnessing the strong familial bonds that exist. Thirdly, there is a need to reduce structural barriers to promoting testing by offering material support to encourage self-isolation ensuring that deprived communities are financially supported. Additionally, leaders or champions from within the community could support and guide through the process of identifying symptoms, accessing testing, explaining results, offering instrumental support and connecting those with positive test results to a healthcare provider for follow-up care. Finally, the stigmatising narrative around EMGs being disproportionately represented for the impact of COVID-19 need to be shifted to acknowledge the complex intersectionality of these issues and the role that deprivation and poverty plays. The media and policy makers should be highlighting the remarkable and enduring resilience of EMGs, many of whom have been over exposed to the virus as they work in critical roles.

## Data Availability

Data is currently unavailable.

## Ethics Approval

The Authors declare that all the research meets the ethical guidelines.

